# Respiratory Tract Pathogen Detection in Children: Clinical Practice and Considerations of mNGS and tNGS Technologies

**DOI:** 10.1101/2025.02.25.25322845

**Authors:** Wang Dujin, Zhan Chuanhua, Yang Hao, Zhu Zhongliang, Zou Yichun

## Abstract

**Objective:** This study aims to evaluate the efficacy of metagenomic next-generation sequencing (mNGS) and targeted next-generation sequencing (tNGS) technologies in detecting respiratory tract pathogens in hospitalized children.

**Methods:** We selected 1275 children hospitalized for respiratory tract infections from March to November 2024 as the study subjects. Based on clinical needs, mNGS/tNGS testing was conducted on these children, including 1225 oropharyngeal swab samples and 50 bronchoalveolar lavage fluid samples. Additionally, traditional microbial culture methods were used for testing in 574 children.

**Results:** The study revealed that children aged 3 to 6 were the primary population affected by respiratory tract infections, accounting for 37.65% of the cases, with 25.1% of them having lower respiratory tract infections. Bacterial and viral co-infections were the most common infection patterns in both oropharyngeal swab and bronchoalveolar lavage fluid samples, with positive rates of 55.02% and 46%, respectively. In bronchoalveolar lavage fluid samples, Mycoplasma pneumoniae had the highest infection rate at 41.67%, followed by Streptococcus pneumoniae and Haemophilus influenzae. The highest detection rate in bronchoalveolar lavage fluid samples was for human adenovirus at 28.3%, followed by rhinovirus and human parainfluenza virus type 3. In oropharyngeal swab samples, Streptococcus pneumoniae had the highest detection rate, followed by Staphylococcus aureus and Haemophilus influenzae. Rhinovirus had the highest detection rate in oropharyngeal swab samples at 25.07%, followed by cytomegalovirus (CMV) and human adenovirus. There was a high consistency between mNGS/tNGS and traditional culture methods in bacterial detection, with the top three being Streptococcus pneumoniae, Staphylococcus aureus, and Haemophilus influenzae. The serum IL-6 concentration in patients with single bacterial infections was higher than in those with single viral infections; the serum IL-6 in patients with bacterial and viral co-infections was higher than in those with single infections, with statistically significant differences. There were statistically significant differences in serum PCT concentration between patients with single bacterial infections and single viral infections, but no significant differences between patients with single bacterial infections and those with bacterial and viral co-infections. The most frequently detected resistance genes included CTX-M-108, NDM-6, OXA-163, tetM, KPC-2, and mecA.

**Conclusion:** mNGS/tNGS technology significantly improved the detection rate of respiratory tract pathogens in children, showing clear advantages over traditional bacterial culture methods in terms of timeliness, accuracy, and coverage of pathogen detection, providing strong technical support for early diagnosis and intervention of respiratory infections in children.

## Introduction

Respiratory infections are among the most common diseases worldwide, posing a significant challenge to public health. These diseases have diverse and complex etiologies, involving numerous pathogens. Therefore, accurate diagnosis of pathogens is crucial for formulating effective treatment plans. Although traditional detection methods, such as smear examination, pathogen culture, biochemical testing, and polymerase chain reaction (PCR), are widely used in clinical settings, they have certain limitations in sensitivity, specificity, and the range of pathogens detected, especially when identifying rare or emerging pathogens [1].

In recent years, the development of high-throughput sequencing technologies, especially next-generation sequencing (NGS), has brought revolutionary progress to the diagnosis of infectious diseases[2–4]. In the context of the COVID-19 pandemic, a global public health event, NGS technology has garnered wider attention and has been widely applied in the detection of pathogens, enabling primary medical institutions to utilize this technology for pathogen detection and identification.

Metagenomic Next Generation Sequencing (mNGS) technology has shown great potential in clinical diagnosis with its high sensitivity and accuracy, capable of rapidly and comprehensively detecting almost all types of pathogens from clinical samples[5]. However, mNGS technology also faces challenges in clinical application, including the complexity of result interpretation, interference from human and background bacterial genomes, and the inability to directly detect antibiotic resistance genes[6–9]

To overcome these limitations of mNGS technology, Targeted Next Generation Sequencing (tNGS) technology has emerged. tNGS technology enriches pathogenic nucleic acids in clinical samples through targeted capture and high-throughput sequencing, then compares and analyzes them with databases to accurately identify pathogens in the samples. tNGS technology not only improves the detection rate of specific pathogens, such as tuberculosis, fungi, and intracellular bacteria, but also detects antibiotic resistance and virulence factor genes, and is more cost-effective than mNGS, facilitating its wide application in clinical infectious disease management[10].

This study aims to explore the application value of mNGS and tNGS technologies in the detection of respiratory tract pathogens and antibiotic resistance genes in hospitalized children, in order to provide more precise diagnostic tools for clinical practice and improve patient treatment outcomes. By comparing with traditional bacterial culture techniques, we hope to provide new perspectives and solutions for the diagnosis and treatment of respiratory infections. Especially during periods of multiple pathogen prevalence, clinical consensus on the diagnosis and treatment of respiratory infections emphasizes the practice and consideration of mNGS and tNGS in the etiological diagnosis of infectious diseases, which provides scientific support for improving the clinical physician’s ability to diagnose and treat respiratory infections and reducing the harm of respiratory infectious diseases.

## Materials and Methods

### 1.1 Study Design and Data Collection

A total of 1275 children hospitalized for respiratory infections from March 2024 to November 2024 were selected as the study subjects. According to the clinical physicians’ needs, 1275 patients underwent mNGS/tNGS testing, including 1225 oropharyngeal swab samples and 50 bronchoalveolar lavage fluid samples. All samples were tested with mNGS or tNGS within 24 hours, and some samples were also tested or not tested with traditional microbial culture methods according to clinical needs.

### 1.2 Ethical approval

The research related to human use has been complied with all the relevant national regulations, institutional policies and in accordance with the tenets of the Helsinki Declaration, and has been approved by the Ethics Committee of Huangshi Central Hospital (NO.2025-4).

### 1.3 Metagenomic Sequencing

Sample nucleic acids were extracted using commercial kits. Pathogen microorganism enrichment was performed using mNGS pathogen enrichment reagents; pathogen-targeted multiple amplification was performed using tNGS pathogen enrichment reagents. Sample library construction was carried out using metagenomic DNA library reagents (reversible terminator sequencing method). Genomic sequencing was performed using the Illumina NextSeq 550 sequencer.

### 1.4 Traditional Microbiological Culture

Microbiological culture was conducted according to clinical needs. The process was carried out in accordance with laboratory standard operating procedures.

### 1.5 Data Statistics

Quantitative data involved in this paper are represented by (N), and positive rates are represented by (%). Statistical analysis was performed using SPSS 25.0 software, and group comparisons were made using χ^2^ tests and t-tests. A P-value of less than 0.05 was considered statistically significant.

## 2. Results

### 2.1. Patient Clinical Characteristics

The study included confirmed that children aged 3 to 6 years old are the majority of respiratory infection patients, accounting for 37.65%. Out of 1275 patients, only two patient samples did not detect any microorganisms. The proportion of male children reached 62.43%. Children with lower respiratory tract infection characteristics accounted for 25.1%, and other fever, cough, and accompanying symptoms are detailed in Table 1.

**Table 1.**
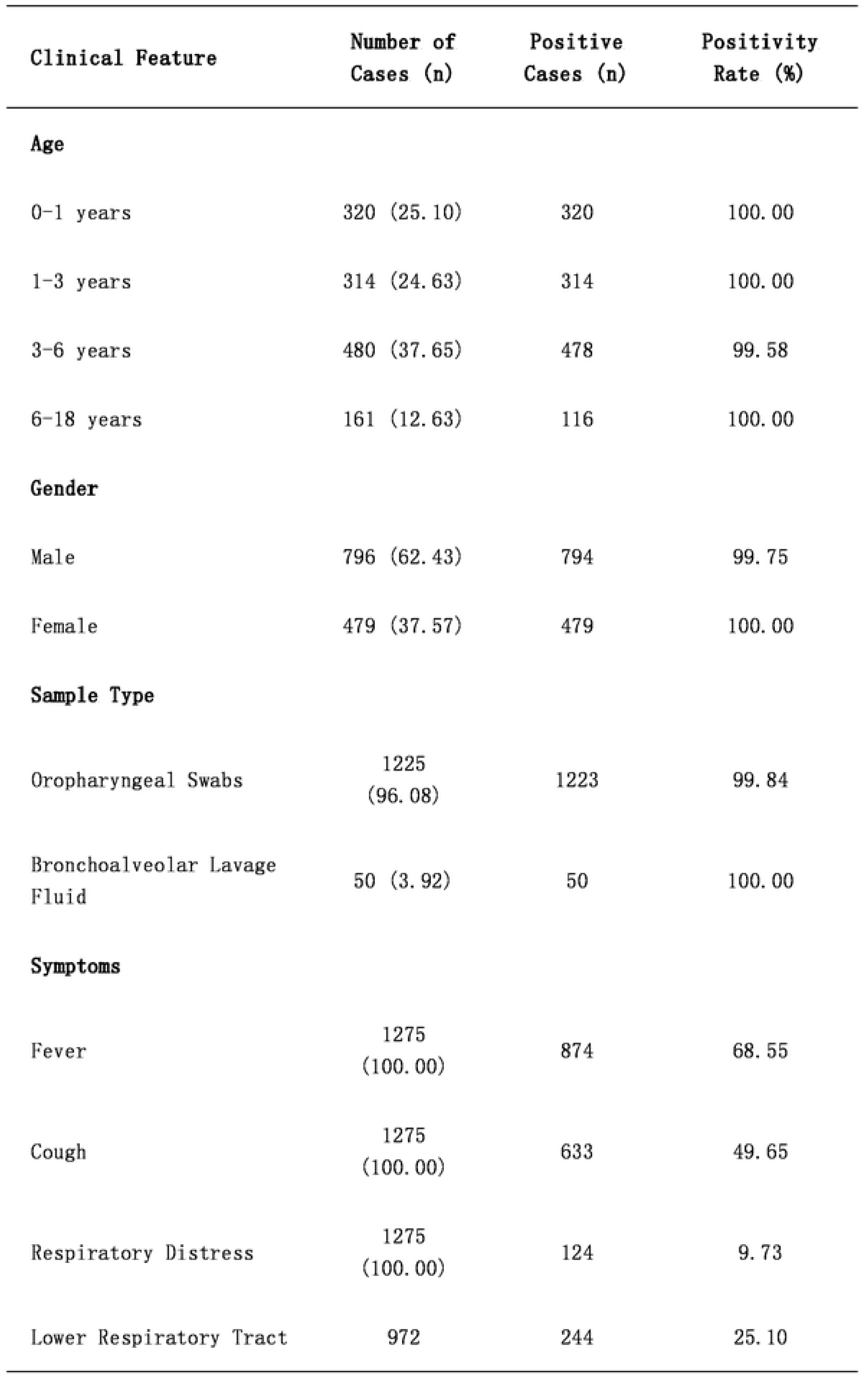
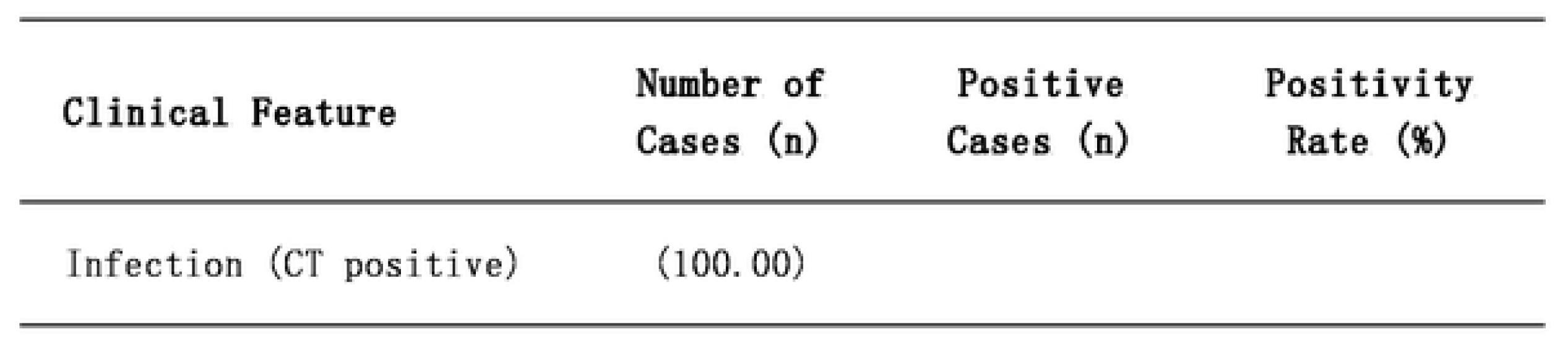
Patient Clinical Characteristics

### 2.2 Analysis of Positive Rates of Single and Multiple Infections Detected by Different Types of Samples with mNGS/tNGS

A total of 1275 patient samples were tested in this study, including 1225 oropharyngeal swab samples and 50 bronchoalveolar lavage fluid samples. Among the oropharyngeal swab samples, the proportion of patients co-infected with bacteria and viruses (B+V) was the highest, reaching 55.02%. Among the bronchoalveolar lavage fluid samples, the proportion of patients co-infected with bacteria and viruses (B+V) was the highest, at 46%.

When comparing oropharyngeal swab and bronchoalveolar lavage fluid samples, there was a statistically significant difference in the positive rates of single bacterial or viral infections. In the comparison of multiple infection positive rates, there was a statistically significant difference in co-infections of bacteria, viruses, and fungi in both types of samples. There was no statistically significant difference between oropharyngeal swab and bronchoalveolar lavage fluid samples in the most common infection pattern (B+V), as detailed in Table 2.

**Table 2.** Positive Rates of Single and Multiple Infections Detected by Different Types of Samples with mNGS/tNGS.

| Group | Number of Samples | B | V | F | B+V | B+F | V+F | B+V+F |
| --- | --- | --- | --- | --- | --- | --- | --- | --- |
| Oropharyngeal Swabs | 1225 | 235(19.18) | 264(21.55) | 3(0.24) | 674(55.02) | 10(0.82) | 11(0.90) | 28(2.29) |
| Bronchoalveolar Lavage Fluid | 50 | 19(38.00) | 1(2.00) | 0(0.00) | 23(46.00) | 1(2.00) | 0(0.00) | 6(12.00) |
| Chi-Square ( $\chi^2$ ) Value | | 10.6600 | 11.1500 | 0.1227 | 1.5770 | 0.7869 | 0.4529 | 17.4700 |
| P Value |  | 0.0011 | 0.0008 | 0.7261 | 0.2092 | 0.3750 | 0.5010 | <0.0001 |

**Notes:**
B: Bacteria
V: Virus
F: Fungus
Chi-square ( $\chi^2$ ) values and P values are used to compare the positivity rates of different pathogen combinations between oropharyngeal swabs and bronchoalveolar lavage fluid samples. A P value < 0.05 indicates a statistically significant difference.

### 2.3 Analysis of Pathogen Detection in Different Samples

All patients included in this study were hospitalized children with respiratory infections, so it is of great significance to comprehensively analyze the infection situation of various types of pathogens. Through overall analysis, the fungal infections detected in this study were mainly Pneumocystis jirovecii, which are categorized as bacteria in Table 3. In terms of bacterial infections, the positive rate of Mycoplasma pneumoniae in bronchoalveolar lavage fluid samples reached 41.67%, followed by Streptococcus pneumoniae and Haemophilus influenzae. In oropharyngeal swab samples, the highest detection rate was Streptococcus pneumoniae, followed by Staphylococcus aureus and Haemophilus influenzae, as detailed in Table 3. In terms of viral infections, the highest detection rate in bronchoalveolar lavage fluid samples was human adenovirus, reaching 28.3%, followed by rhinovirus and human parainfluenza virus type 3. In oropharyngeal swab samples, the highest detection rate was rhinovirus, reaching 25.07%, followed by human cytomegalovirus (CMV) and human adenovirus, as detailed in Table 4.

**Table 3.**
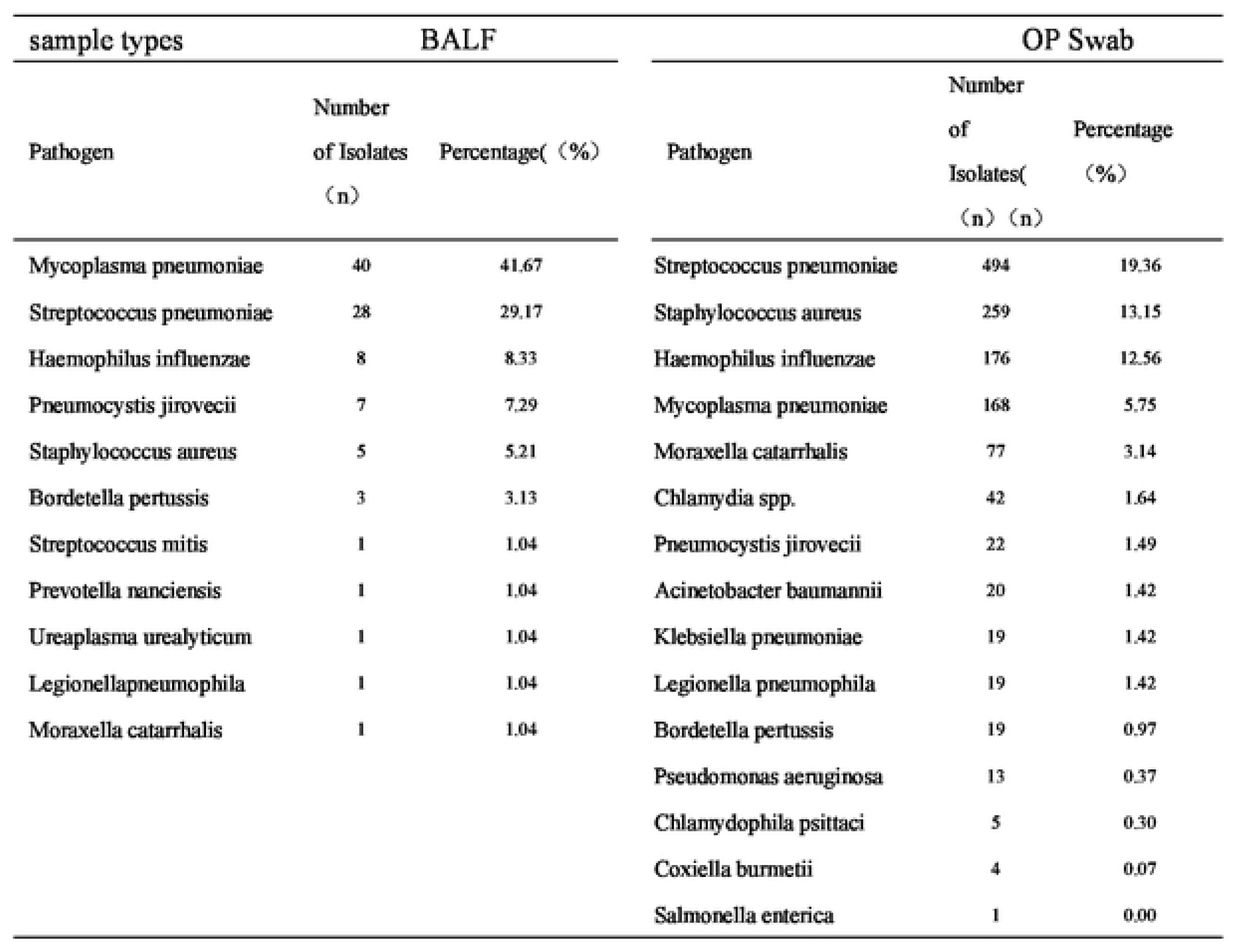
Bacterial Pathogen Detection in Different Samples.

**Table 4.**
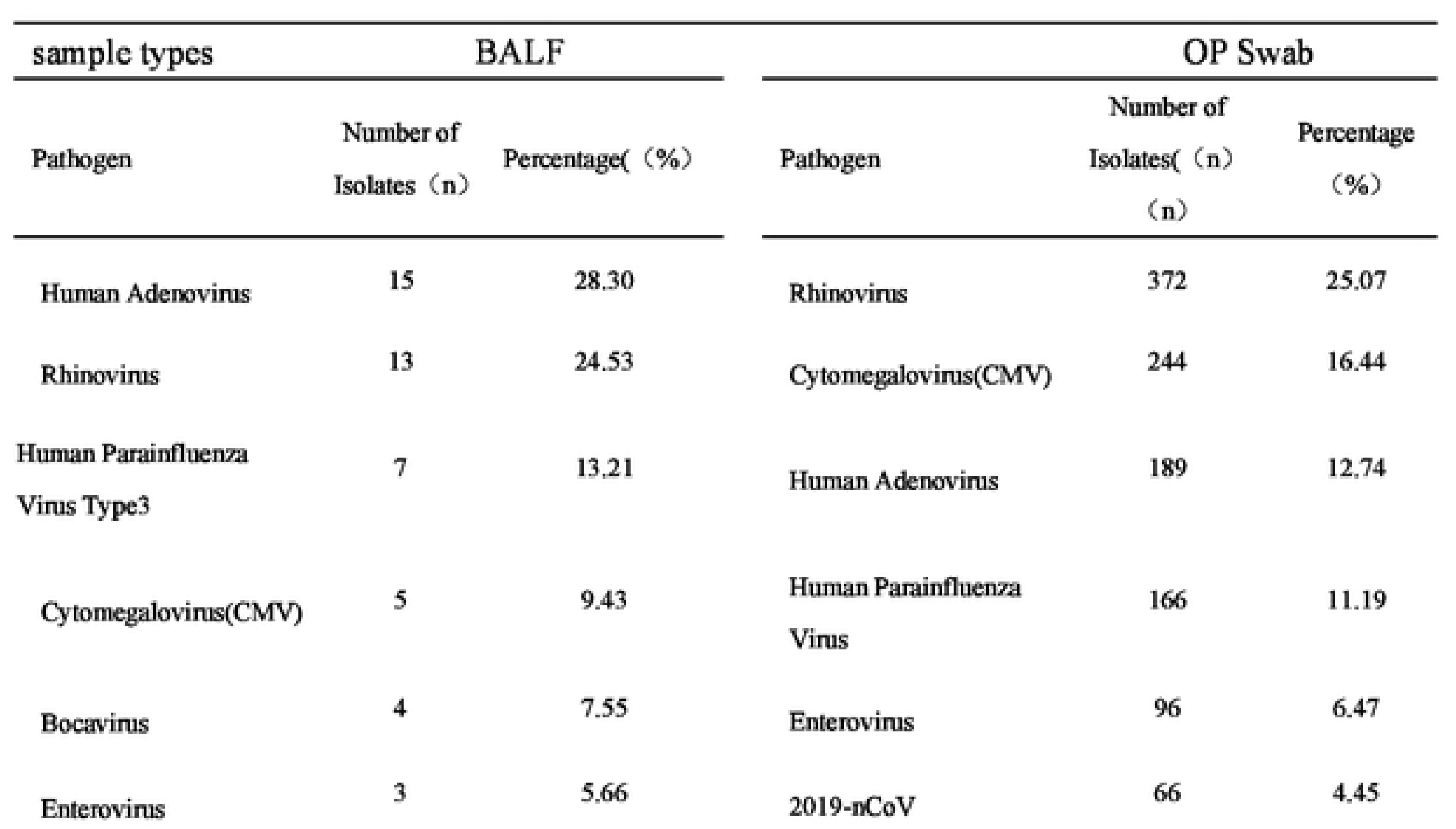
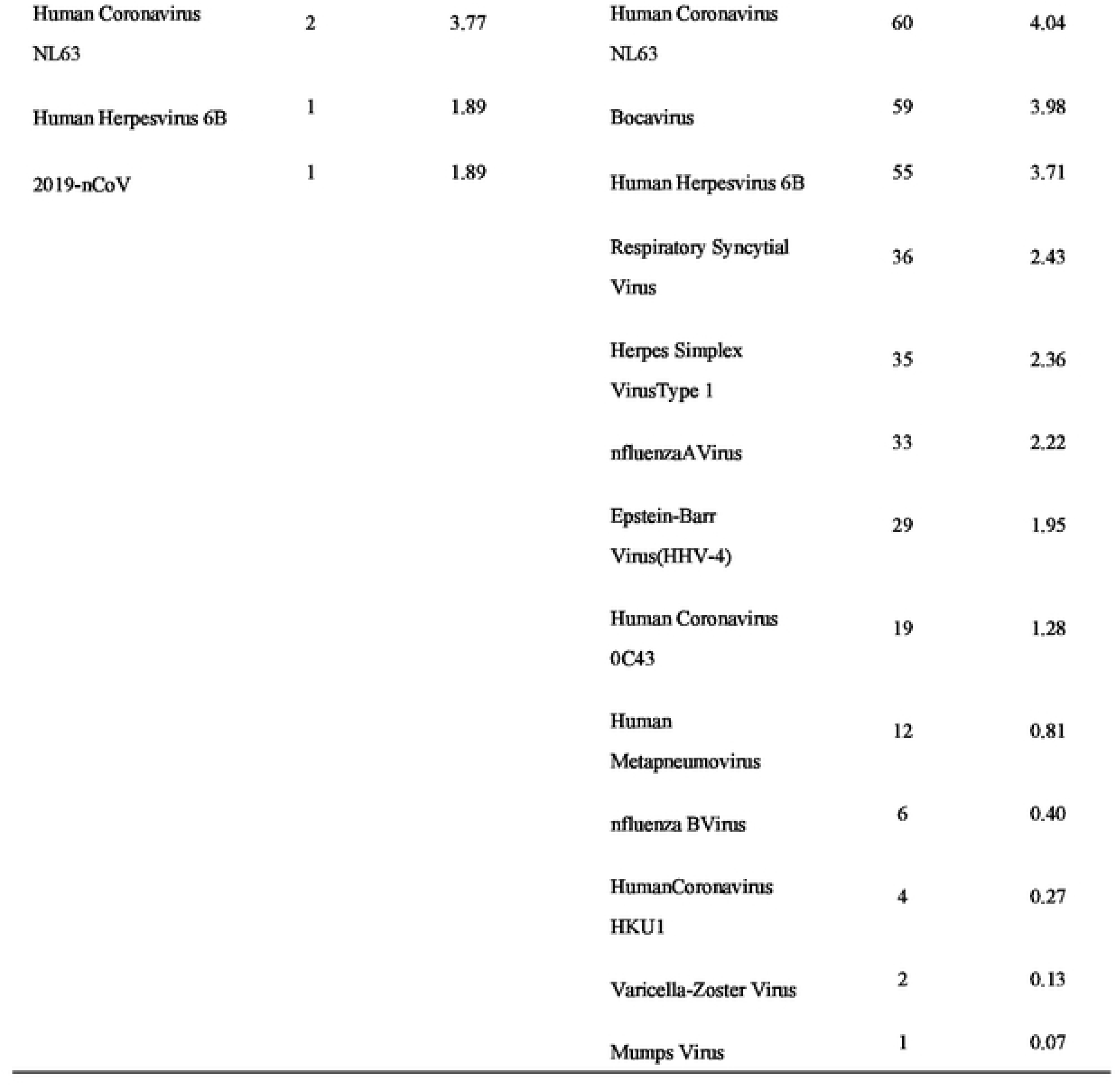
Viral Pathogen Detection in Different Samples.

### 2.4 Comparison of Positive Results between mNGS/tNGS and Microbiological Culture Methods

A total of 1275 samples were included in this study, with 1225 oropharyngeal swab samples and 50 bronchoalveolar lavage fluid samples. Microbiological culture was performed on 537 samples according to the clinical physicians’ needs. Limited to the principles of microbiological detection methods, only positive bacteria were compared here. Among them, 99 cases were positive for the same type of bacteria in both mNGS/tNGS and microbiological culture methods, accounting for 18.44%; 13 cases were not positive for bacteria in both mNGS/tNGS and microbiological culture methods, accounting for 2.23%; 12 cases were positive for bacteria in microbiological culture methods but not the same type of bacteria in mNGS/tNGS, accounting for 14%; 343 cases were positive for bacteria in mNGS/tNGS but not in culture methods, accounting for 63.87%, as detailed in Table 5. The bacteria detected most by mNGS/tNGS and the bacteria detected most by traditional culture methods were highly consistent, with the top three bacteria being Streptococcus pneumoniae, Staphylococcus aureus, and Haemophilus influenzae, as shown in Figure 1.

**Figure 1.**
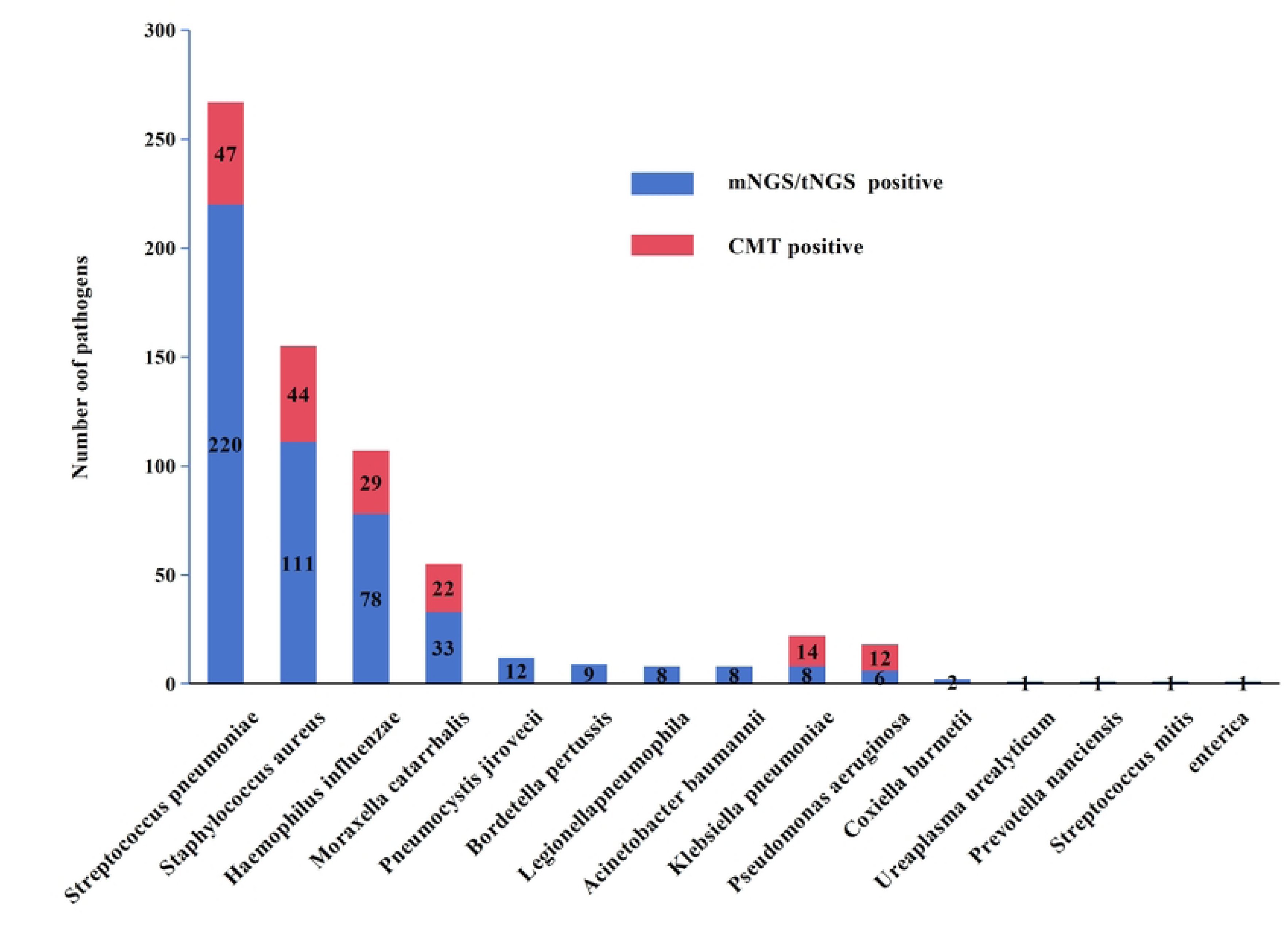
Comparison of Positive Results between mNGS/tNGS and Microbiological Culture Methods

**Figure 2.**
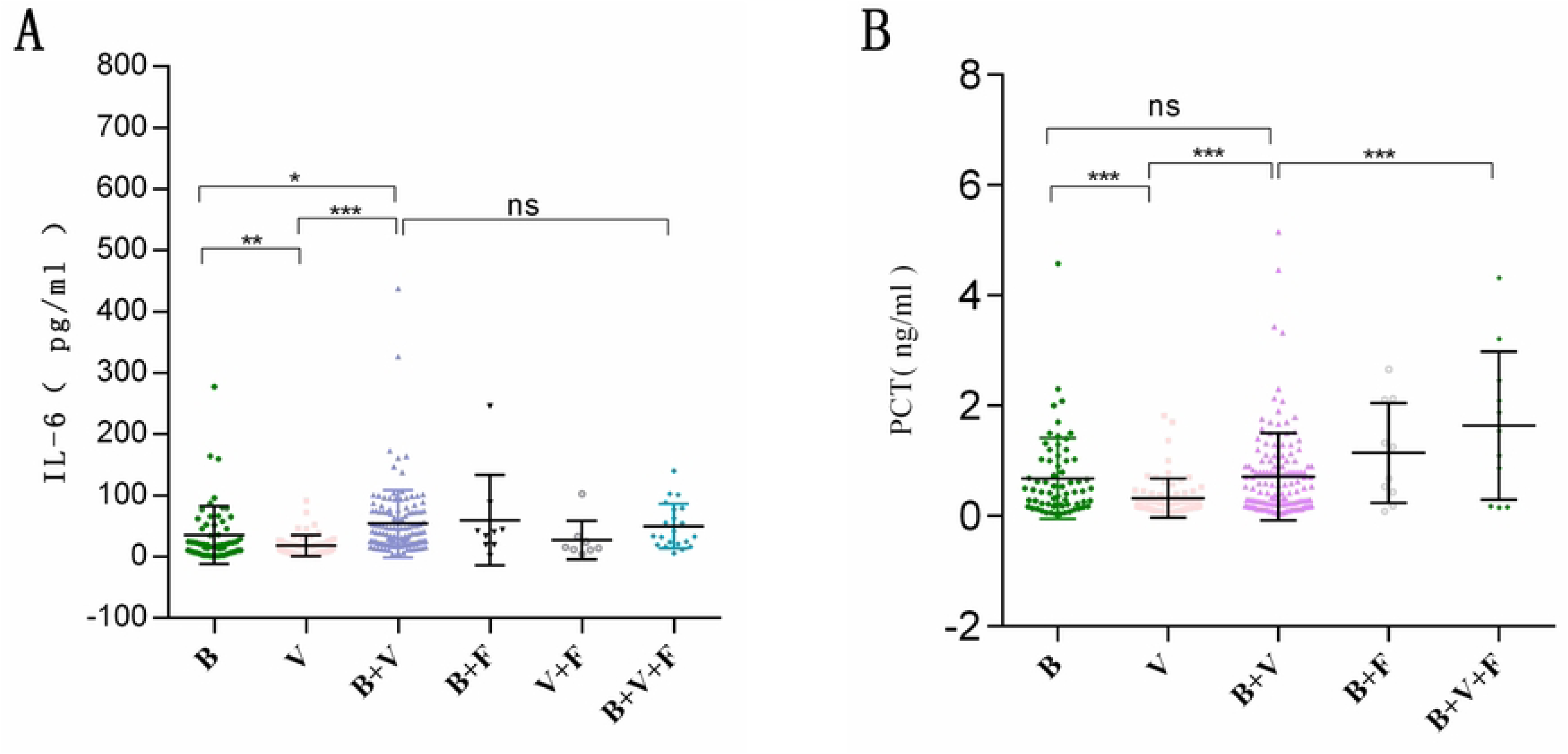
Detection of Serum Inflammatory Markers

**Table 5.** Comparison of the positive rate of bacteria detected by traditional microbial culture method and mNGS/tNGS.

| CMT | mNGS/tNGS |  |  | P Value |
| --- | --- | --- | --- | --- |
|  | Positive | Negative | Total |  |
| Positive | 99 | 83 | 182 | <0.0001 |
| Negative | 343 | 12 | 355 |  |
| Total | 442 | 95 | 537 |  |

\*Use Fisher's exact test.

### 2.5 Detection of Serum Inflammatory Markers

This study focused on the correlation between different types of pathogen infections and two infection markers, Procalcitonin (PCT) and Interleukin-6 (IL-6). The study found that there was a statistically significant difference in serum IL-6 concentrations between patients with single bacterial infections and those with single viral infections, with IL-6 concentrations being higher in patients with single bacterial infections. Patients with mixed bacterial and viral infections had higher serum IL-6 levels than those with single bacterial or viral infections, which was statistically significant. Notably, there was no statistically significant difference in IL-6 concentrations between patients with dual bacterial and viral infections and those with triple infections involving bacteria, viruses, and fungi. The study indicated that patients with single bacterial infections had higher serum PCT concentrations than those with single viral infections, which was statistically significant. However, there was no statistically significant difference in PCT levels between patients with single bacterial infections and those with mixed bacterial and viral infections, suggesting that bacteria are the primary cause of elevated PCT in patients with respiratory infections. Additionally, patients with triple infections involving bacteria, viruses, and fungi had higher serum PCT levels than those with dual bacterial and viral infections, which was statistically significant, indicating that fungal infections can promote an increase in PCT serum levels, and PCT shows higher specificity in distinguishing different types of infections, especially in bacterial infections.

### 2.6 Detection of Antibiotic Resistance Genes

Patients who underwent tNGS testing were also tested for related antibiotic resistance genes. Out of 1225 patients, the most frequently detected resistance gene was CTX, with 196 cases. The most common subtype of CTX was CTX-M-108, with 69 cases, accounting for 35.2%; Tet resistance genes were detected in 83 cases, with the most common subtype being tetM, with 45 cases, accounting for 54.22%; KPC resistance genes were detected in 188 cases, with the most common subtype being KPC-2, with 179 cases, accounting for 95.21%; OXA resistance genes were detected in 28 cases, with the most common subtype being OXA-163, with 14 cases, accounting for 50%; NDM resistance genes were detected in 51 cases, with the most common subtype being NDM-6, with 39 cases, accounting for 76.47%; other resistance genes were detected in 99 cases, with the most common subtype being mecA, with 45 cases, accounting for 45.45%. See Figure 3 for details.

**Figure 3.**
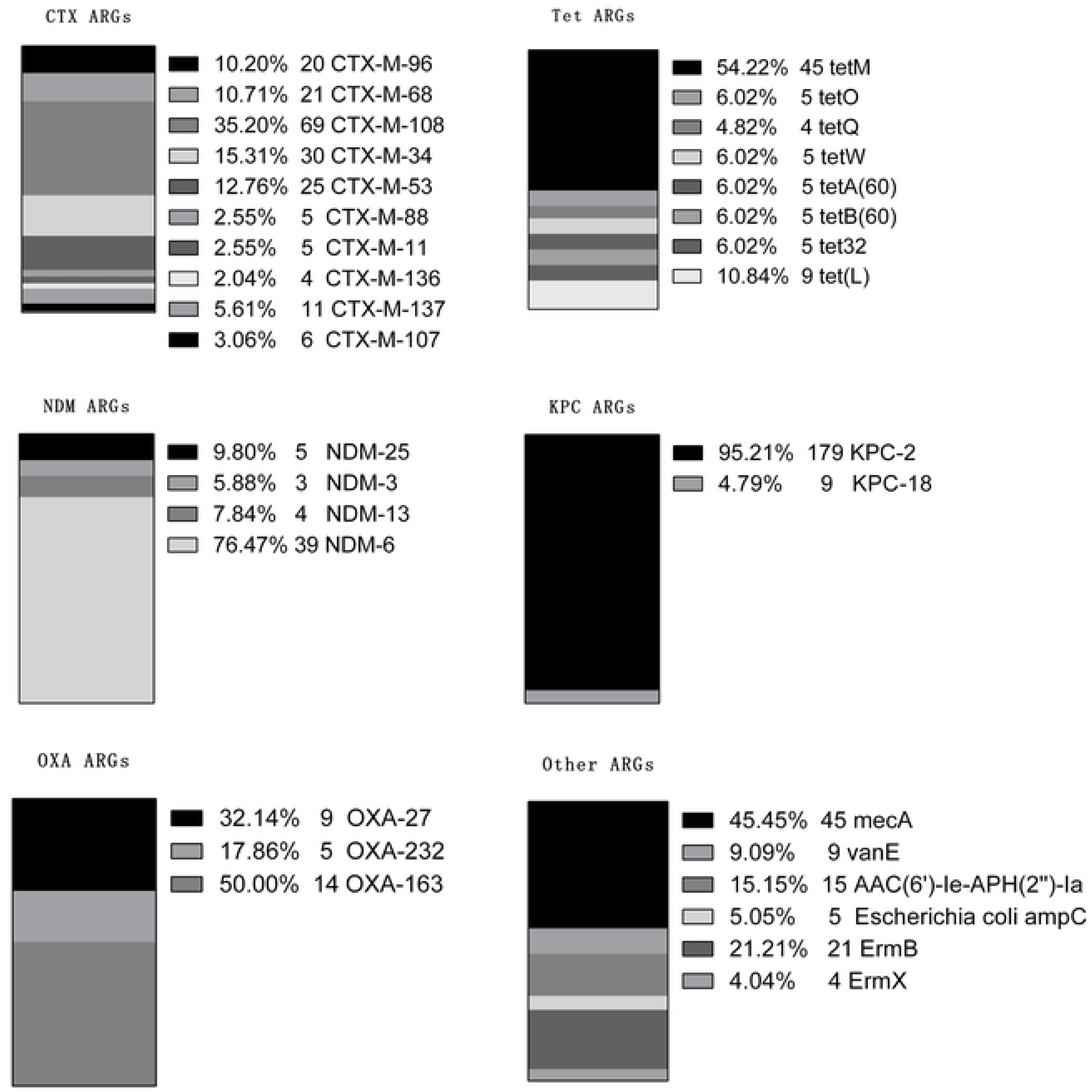
Detection of Antibiotic Resistance Genes

## 3. Discussion

This study conducted tNGS or mNGS testing on 1275 hospitalized children and found that children aged 3 to 6 years old accounted for the highest proportion, reaching 37.65%. This finding is consistent with recent studies on the epidemiology of respiratory infections in children, emphasizing the high risk of this age group for respiratory infections [11]. Among the patients included in the study, boys accounted for 62.43%. In terms of sample types, oropharyngeal swabs were the majority, likely due to their non-invasive collection method, which is easier to implement in children [12]. Additionally, we found that 25.1% of patients developed lower respiratory tract infections, a proportion that highlights the need for closer monitoring and timely treatment for this group of patients [13].

Our study results emphasize the significant advantage of high-throughput sequencing technology in terms of sensitivity and coverage compared to traditional microbiological culture methods. The combined use of tNGS/mNGS and microbiological culture techniques increased the positive detection rate of bacteria to 97.77% (525/537), far exceeding the 18.44% (99/537) obtained by using microbiological culture methods alone. This finding echoes recent studies that also emphasize the increased accuracy of pathogen detection when these two technologies are used in combination [14–15].

In this study, most respiratory infections in hospitalized children were mixed infections, a phenomenon consistent with the global pattern of respiratory infections [16]. We found that in both oropharyngeal swab and bronchoalveolar lavage fluid samples, the most common type of mixed infection was bacteria plus viruses (B+V), accounting for 55.02% and 46% respectively. This may be related to the ability of viruses to damage the epithelial barrier, making it easier for bacteria to invade and colonize [17].

In this study, there was a significant difference in the types of positive pathogens between bronchoalveolar lavage fluid samples and oropharyngeal swab samples, which may be related to the different sampling sites. Bronchoalveolar lavage fluid can more directly reflect the pathogen situation in the lower respiratory tract [18–19]. In terms of bacterial detection rates, the positive rate of Mycoplasma pneumoniae in bronchoalveolar lavage fluid samples was much higher than in oropharyngeal swab samples, which is consistent with the clinical observation that Mycoplasma pneumoniae mainly causes lower respiratory tract infections [20]. In oropharyngeal swab samples, the highest detection rate was for Streptococcus pneumoniae. In terms of viral detection rates, human adenovirus was the most prevalent in bronchoalveolar lavage fluid samples, while rhinovirus was the most prevalent in oropharyngeal swab samples, which is in line with the recent trend of rhinovirus prevalence [21].

Serum inflammatory markers IL-6 and PCT are of great value in assessing infection status. This study found that patients with mixed infections of bacteria and viruses or fungi had higher IL-6 expression levels in serum than those with single infections, which is consistent with the key role of IL-6 in the inflammatory response [22]. However, there was no statistically significant difference in IL-6 concentrations between patients with dual bacterial and viral infections and those with triple infections involving bacteria, viruses, and fungi. Patients with single bacterial infections had higher serum PCT concentrations than those with single viral infections, which was statistically significant. However, there was no statistically significant difference in PCT levels between patients with single bacterial infections and those with mixed bacterial and viral infections, indicating that bacteria are the main cause of elevated PCT in patients with respiratory infections. Additionally, patients with triple infections involving bacteria, viruses, and fungi had higher serum PCT levels than those with dual bacterial and viral infections, which was statistically significant, indicating that fungal infections can promote an increase in PCT serum levels, and PCT shows higher specificity in distinguishing different types of infections, especially in bacterial infections [23].

The detection of antibiotic resistance genes provides important basis for personalized treatment for clinicians. The detection of resistance genes such as CTX-M-108 and KPC-2 in this study provides guidance for the rational use of antibiotics [24]. Nevertheless, tNGS/mNGS technology still faces challenges in clinical application, including the complexity of the detection process, interference from human and background bacterial genomes, and the difficulty in distinguishing between colonizing bacteria and opportunistic pathogens [25]. These challenges require close communication between laboratory physicians and clinicians to ensure the clinical relevance of the test results [26].

## Data Availability

All relevant data are within the manuscript and its Supporting Information files.

## Acknowledgments

We would like to thank all the patients and their families who participated in this study. We also thank the clinical and laboratory staff at Huangshi Central Hospital for their support and assistance.

## Ethics Approval and Consent to Participate

The study was conducted in accordance with the declaration of Helsinki. This study was approved by the Medical Ethics Committee of Huangshi Central Hospital, Affiliated Hospital of Hubei Polytechnic University. This clinical study is a retrospective study that only collects patient clinical data and does not interfere with the patient’s treatment plan. It does not pose any physiological risks to the patient. Patient’s privacy will be respected in accordance with the Declaration of Helsinki.

## Author Contributions

Zou Yichun conceived and designed the study.Zhan Chuanhua and Yang Hao collected the samples and performed the experiments.Zhu Zhongliang analyzed the data. Wang Dujin wrote the manuscript. All authors reviewed and approved the final version of the manuscript.

## Conflict of Interest Statement

The authors declare that they have no competing interests.

## Funding

This study was supported by Hubei Key Laboratory of Kidney Disease Pathogenesis and Intervention (Grant Number:SB202111).

## Notes

### Competing Interest Statement

The authors have declared no competing interest.

### Funding Statement

The author(s) received no specific funding for this work.

